# Effects of repetitive transcranial magnetic stimulation on contralesional dorsal premotor cortex in subcortical stroke patients with moderate to severe upper limb motor impairment: a pilot randomized controlled trial

**DOI:** 10.1101/2025.03.05.25323209

**Authors:** Shan Lu, Hewei Wang, Yujian Yuan, Tianyu Ma, Zhichao Liu, Qiguang Li, Yunhui Fan, Jingjing Zhu, Zhaoying Ji, Yun Chen, Tianhao Gao, Qing Xu, Limin Sun

**Affiliations:** Department of Rehabilitation Medicine, Huashan Hospital Fudan University, Shanghai, China; Department of Rehabilitation Medicine, Shanghai NO.3 Rehabilitation Hospital, Shanghai, China

**Author notes:** Correspondence authors: **Limin Sun**, Department of Rehabilitation Medicine, Huashan Hospital Fudan University, Shanghai, China; **Qing Xu**, Department of Rehabilitation Medicine, Shanghai NO.3 Rehabilitation Hospital, Shanghai, China. These authors contributed equally to this work (Shan Lu, Hewei Wang).

**Keywords:** Transcranial magnetic stimulation, Dorsal premotor cortex, Primary motor cortex, Stroke, Upper limb

## Abstract

**Objective:** This randomized controlled study examines the effect of repetitive transcranial magnetic stimulation on contralesional dorsal premotor cortex in patients in the subaucte phase of stroke recovery.

**Methods:** This randomized controlled study includes three groups: contralesional dorsal premotor cortex(cPMd), contralesional primary motor cortex(cM1) and sham stimulation. 60 Participants were divided into 3 groups according to random block method. They all received a 3-week intervention. The primary outcome was using the Fugl Meyer Assessment (FMA) to evaluate the motor function of the affected limb. The secondary outcomes included: Brunnstrom hemiplegia grading, Action Research Arm Test (ARAT), Wolf Motor Function Test (WMFT), Barthel Index (BI), electrophysiological assessment (bilateral hemisphere MEP, ipsilesional central conduction time, ipsilesional latency).

**Results:** Comparative analysis revealed that the cPMd group demonstrated significant improvements in both the Fugl-Meyer Assessment for Upper Extremity (FMA-UE) and distal upper limb scores relative to the cM1 group (P<0.05) and the sham group (P<0.001). What’s more, the cPMd group exhibited marked enhancementsin FMA for Lower Extremity (FMA-LE), proximal upper limb, and hand scores (P≤0.01) compared to the sham stimulation group. However, the study revealed limited improvements in electrophysiological assessments, with only ipsilesional central conduction time showing significant changes compared to the sham group.

**Discussion:** This study provides some evidence for the effect of high-frequency rTMS stimulation in the cPMd on improving stroke motor disorders. The cPMd group showed significant improvement in FMA-UE scores compared to the cM1 group. It suggests that high-frequency rTMS in the cPMd may be a unique and more effective strategy for optimizing brain motor networks and improving patient dysfunction. This provides new ideas for TMS assessment and intervention of brain areas based on complex brain networks and proposes new possible treatment strategies. In the future, a multi-center, large-scale systematic study is needed to explore the use of high-frequency rTMS in the cPMd for stroke hemiplegia recovery, and to further investigate the pathophysiological mechanisms of disease recovery, as well as to explore the neural plasticity mechanism of stroke motor dysfunction recovery. Therefore, the rehabilitation potential of high-frequency rTMS in the cPMd still needs further exploration and better promotion of motor function recovery in stroke patients.

## Introduction

Stroke is a leading cause of global mortality and long-term disability, with persistent motor dysfunction affecting a significant proportion of survivors despite some degree of spontaneous recovery[1]. The recovery of motor function in patients with chronic stroke is a difficult point, and most patients are still unable to complete simple daily activities with their affected hands after 6 months of illness[2]. Repetitive transcranial magnetic stimulation (rTMS), a non-invasive neuromodulation technique, has emerged as a promising intervention for enhancing motor recovery in stroke patients by modulating cortical excitability. High-frequency rTMS stimulation can enhance cortical excitability, while low-frequency rTMS can reduce cortical excitability and increase contralateral excitability. For stroke patients, low-frequency stimulation of the primary motor (M1) cortex in the contralesional hemisphere is widely used to promote motor function recovery after hemiplegia[3]. However, in recent years, there has been no consistent conclusion on the clinical efficacy of rTMS in treating post-stroke hemiplegia. A systematic review indicates that current evidence does not sufficiently support the efficacy of rTMS in conventional stroke treatment[4]. Conversely, a meta-analysis highlights that rTMS may positively influence motor function recovery, particularly during the subacute phase.[5]

The discrepancies in these findings may be attributed to three primary factors: (1) Significant heterogeneity in patient characteristics, including stroke onset time, location, and severity; (2) Many studies rely heavily on the interhemispheric inhibition (IHI) model, which posits that stroke disrupts the inhibitory balance between hemispheres, with the contralesional hemisphere exerting inhibitory effects on the ipsilesional cortex, thereby exacerbating functional impairment of the affected limb[6]; (3) Guided by the IHI model, current rTMS protocols primarily involve low-frequency stimulation of the contralesional M1 cortex to reduce excitability in the contralesional hemisphere and mitigate interhemispheric imbalance.

Many recent studies have suggested that the contralesional motor cortex has a promoting effect on the motor function of hemiplegic limbs, especially in patients with severe injuries on the affected side, which contradicts the IHI model[7, 8]. Therefore, some people have proposed a bimodal balance–recovery model for stroke, suggesting that the role of the contralesional motor cortex in the affected side’s motor function may vary depending on the degree of preservation of normal structures on the affected side: inhibition is dominant when more normal structures are preserved on the affected side; When there is less preservation of normal structure, compensation is the main approach[9]. Therefore, the boundary point of structural preservation may be helpful in determining the treatment plan for rTMS, but there are no literature reports on the boundary values of related brain connectivity parameters.

Based on the hemisphere compensation theory in the bimodal balance–recovery model, high-frequency rTMS treatment in the contralesional hemisphere may achieve better results for patients with less preservation of normal structures, but there is no definite conclusion on the target brain area in the contralesional hemisphere. The contralesional dorsal premotor cortex (cPMd) in the secondary motor cortex is a potential target[10, 11]. During hand movement on the affected side, higher activation can be observed in the cPMd cortex, which is directly proportional to the degree of stroke injury. The preliminary research found that the mediating centrality of cPMd was significantly reduced compared to the healthy control group, and was positively correlated with the motor function score of the affected upper limb. It is speculated that high-frequency stimulation of cPMd in patients can promote functional recovery through the healthy side compensation model[12]. Current research gaps include limited focus on cPMd[8, 13, 14], insufficient longitudinal data on standardized intervention protocols. This study aims to clarify the therapeutic effect of high-frequency rTMS stimulation in the cPMd on stroke patients with motor disorders and compare its efficacy with traditional low-frequency rTMS stimulation intervention in the cM1. The study aims to validate, based on the dual-mode balance recovery model, whether high-frequency repetitive transcranial magnetic stimulation (HF-rTMS) applied to the cPMd demonstrates superior efficacy compared to low-frequency rTMS (LF-rTMS) applied to the cM1 in patients with severe hemiplegic stroke.

## Methods

### Participants

This single-blind, randomized controlled trial (RCT) enrolled 60 patients with subcortical stroke-induced hemiparesis patients admitted to Huashan Hospital of Fudan University and the Third Rehabilitation Hospital of Shanghai from October 2021 to December 2022. Written informed consent was obtained from all participants prior to enrollment. The study protocol was reviewed and approved by the ethics committees of Huashan Hospital of Fudan University and the Third Rehabilitation Hospital of Shanghai.

### Inclusion and Exclusion Criteria

Inclusion criteria were as follows:(1) First-ever stroke; (2) Subcortical stroke confirmed by head CT or MRI; (3) Alert and stable vital signs; (4) Age between 18 and 80 years; (5) Disease duration of 3 to 12 months; (6) Hemiparesis with Brunnstrom grade ≤ IV in the affected hand; (7) Right-handedness.

Exclusion criteria included: (1) Severe communication, attention, hearing, vision, sensory, intellectual, psychiatric, or cognitive impairments (Mini-Mental State Examination score < 27); (2) Severe spasticity or pain; (3) Musculoskeletal, neurological, or other systemic diseases, malignant tumors, or severe cardiopulmonary, hepatic, or renal dysfunction; (4) Inability to complete local follow-up; (5) Presence of metal implants, magnets, or pacemakers; (6) Alcohol or drug dependence; (7) History of epilepsy.

### Randomization

Stroke patients with hemiplegia who met the inclusion criteria were divided into 3 groups according to random block method, with 20 cases in each group, which were cM1 low-frequency group, cPMd high-frequency group and sham stimulation group(sham group), respectively. There was no statistical significance in the comparison of general data among the three groups (P>0.05), which was comparable, as shown in Table 1.

**Table 1.** Basic Information of Three Groups of Patients.

| basic feature | cPmd group<br>n=19 | M1 group<br>n=19 | sham group<br>n=19 | F/X<sup>2</sup> | P |
| --- | --- | --- | --- | --- | --- |
| Age<br>(Year, $\bar{x} \pm SD$) | 64.47 ± 9.01 | 65.37 ± 6.09 | 65.84 ± 6.90 | -.* | 0.994 |
| Gender<br>(Male/Female) | 14/5 | 14/5 | 14/5 | -.* | 1 |
| Types of stroke<br>(Cerebral infarction/bleeding) | 16/3 | 17/2 | 17/2 | -.* | 0.852 |
| course of disease<br>(Month, $\bar{x} \pm SD$) | 4.65 ± 2.20 | 4.05 ± 1.39 | 3.80 ± 1.96 | 1.073 | 0.349 |

\* Due to significant deviations from normality ( $p < 0.05$ ), the Kruskal-Wallis H test was used for between-group comparisons.

### Intervention measures

All three patient groups underwent routine rehabilitation therapy, including physical therapy (PT) for limb motor function training and occupational therapy (OT) for daily living ability training. These sessions were conducted 5 times per week, with each session lasting 40 minutes, over a period of 3 weeks. Concurrently, patients received standard pharmacological treatment. Additionally, all patients received identical traditional therapies, such as biofeedback electrical stimulation, acupuncture, and massage, which were administered over a 3-week period. The cM1 low-frequency group received 1Hz rTMS stimulation targeting the contralesional M1 cortex, with a stimulation intensity set at 90% of the resting motor threshold (RMT). Each session consisted of 10 seconds of stimulation (10 pulses), followed by a 2-second interval, totaling 100 sessions and 1000 pulses per day. This treatment was administered once daily, 5 days per week, with each session lasting 20 minutes. The cPMd high-frequency group received 5Hz rTMS stimulation targeting the PMd region of the healthy hemisphere, located 2 cm anterior and 1 cm medial to the optimal M1 stimulation point[15]. The stimulation intensity was similarly set at 90% RMT. Each session consisted of 10 seconds of stimulation (50 pulses), followed by a 50-second interval, totaling 20 sessions and 1000 pulses per day. This treatment was administered once daily, 5 days per week, with each session lasting 20 minutes. The sham stimulation group underwent the same protocol as the cM1 low-frequency group, with the coil positioned vertically on the skull surface to ensure no effective stimulation was delivered. Based on the aforementioned treatment parameters, all three groups received rTMS therapy for 3 weeks, totaling 15 treatment sessions.

### The outcomes

Clinical functional and electrophysiological assessments were conducted at three time points: baseline (enrollment), 3 weeks post-intervention, and 4 weeks after the completion of the intervention. The primary outcome measure was motor function of the affected limb, assessed using the Fugl-Meyer Assessment (FMA). Secondary outcome measures included: motor function and muscle tone changes of the affected hand, evaluated using the Brunnstrom hemiplegia grading scale; motor function of the affected hand, assessed via the Action Research Arm Test (ARAT); motor function of the affected upper limb, measured using the Wolf Motor Function Test (WMFT); and activities of daily living (ADL) and related abilities, evaluated through the Barthel Index (BI). Additionally, electrophysiological assessments were performed, including motor evoked potentials (MEPs) in both hemispheres, ipsilesional central conduction time, and ipsilesional latency.

### Statistical Analysis

SPSS 24.0 was used for statistical analysis. The Shapiro-Wilk test was employed to assess the normality of data distribution. Continuous variables are presented as mean ± standard deviation (mean ± SD), while categorical variables are expressed as percentages (%). Intergroup comparisons were conducted using one-way ANOVA or the Kruskal-Wallis H test, depending on the normality of the data. Within-group changes from baseline to post-intervention were analyzed using paired t-tests or Wilcoxon signed-rank tests, based on the normality of difference scores as determined by Shapiro-Wilk tests.

## Results

### 1. Baseline Characteristics of the Study Groups

No statistically significant differences were observed in the baseline characteristics among the three groups (p > 0.05), confirming their comparability, as detailed in Table 1. Among the 60 patients (3 patients dropped out), 7 had cerebral hemorrhage, and 50 had cerebral infarction. Baseline assessments revealed no significant differences (P > 0.05) in MMSE, FMA, Brunnstrom hand function staging, WMFT, or ARAT scores among the three groups, further confirming their comparability, as presented in Table 2. No significant adverse reactions were observed during the treatment period across the three groups.

**Table 2.** Baseline Assessment Data of Three Groups of Patients.

| P | cPMd group<br>(n=19) | M1 group<br>(n=19) | sham group<br>(n=19) | P |
| --- | --- | --- | --- | --- |
| MMSE | 27.65 ± 1.87 | 26.7 ± 2.13 | 26.25 ± 1.91 | 0.082 |
| FMA-UE | 27.16 ± 10.13 | 28.42 ± 11.64 | 24.53 ± 10.09 | 0.520 |
| FMA-LE | 22.32 ± 3.53 | 20.89 ± 4.07 | 21.00 ± 5.31 | 0.539 |
| Brunnstrom Hand<br>Function Staging | 2 (2, 3) | 3 (2, 4) | 2 (1, 3) | 0.436 |
| WMFT | 8.95 ± 9.19 | 13.37 ± 11.09 | 9.16 ± 7.11 | 0.439 |
| ARAT | 8.65 ± 9.03 | 13.65 ± 10.86 | 6.69 ± 6.97 | 0.310 |

### 2. Comparison of Brunnstrom Hand Function Stages Before and After Treatment

Both the cPMd and cM1 groups demonstrated significant improvements in Brunnstrom hand function staging post-intervention (P < 0.05). However, no significant differences were observed between the cPMd and cM1 groups compared to the sham group, as detailed in Table 3.

**Table 3.** Comparison of Bromstorm Hand Function Stages Before and After Treatment.

| Group | Before treatment | After treatment |
| --- | --- | --- |
| cPMd group(n=19) | 2 (2, 3) | 3 (2, 3.75) <sup>a</sup> |
| cM1 group(n=19) | 3 (2, 4) | 3.5 (2, 4) <sup>a</sup> |
| sham group (n=19) | 2 (1, 3) | 2 (2, 4) |

<sup>a</sup>P < 0.05, indicates a statistically significant difference compared to before treatment within the group;
<sup>b</sup>P < 0.05, indicates a statistically significant difference compared to the sham group;
<sup>c</sup>P < 0.05, indicates a statistically significant difference compared to the cM1 group.

### 3. Comparison of FMA-UE Scores Before and After Treatment

FMA-UE scores significantly improved in all three groups post-treatment (P<0.05). The cPMd group exhibited significantly greater improvements in FMA-UE scores compared to the sham group (P < 0.05). The cPMd group demonstrated the highest degree of improvement in FMA-UE scores compared to the other groups (<0.05), while the cM1 group showed greater improvement than the sham group, though this difference was not statistically significant (P = 0.065), as detailed in Table 4.

**Table 4:**
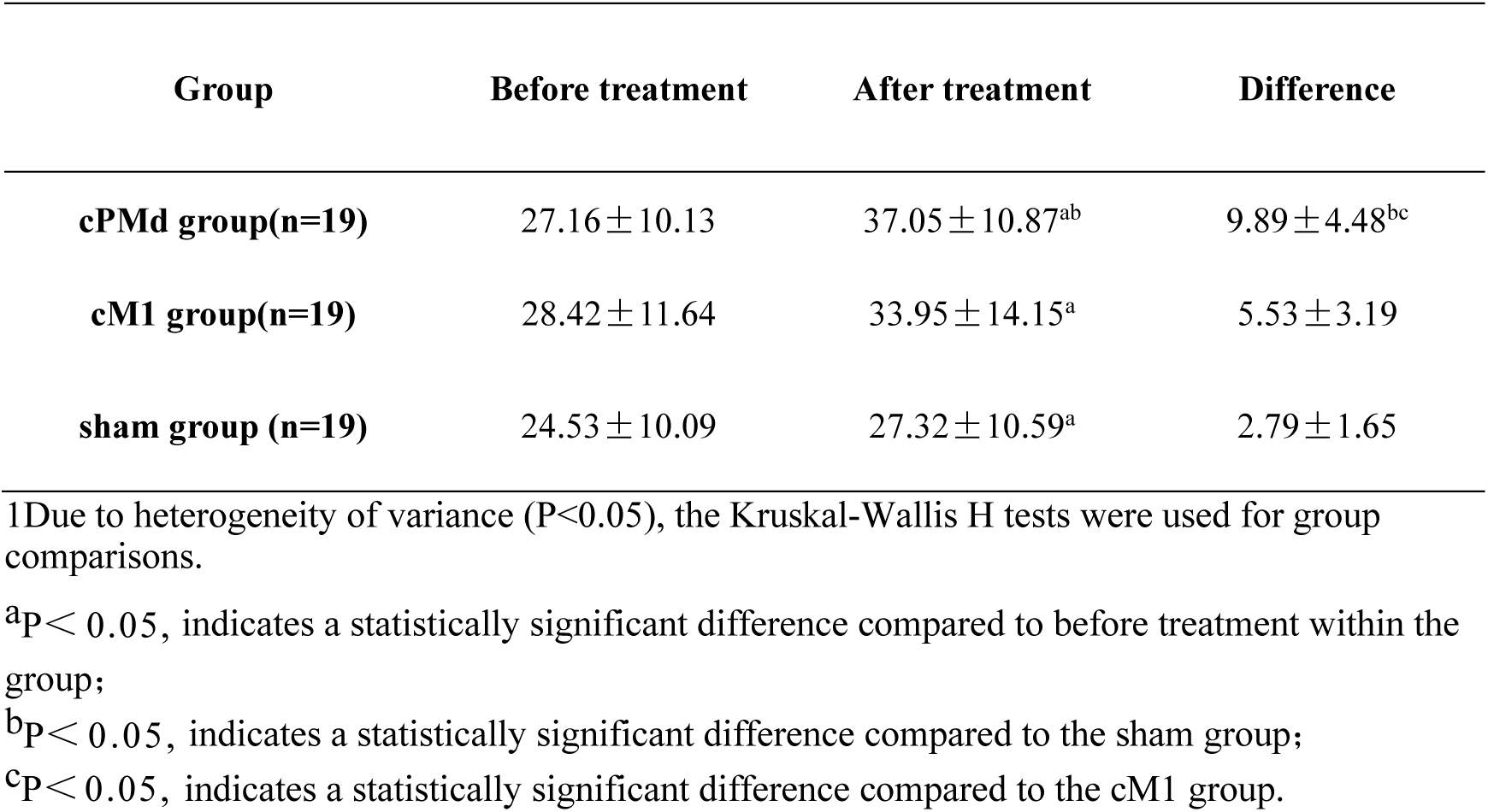
Comparison of FMA-UE Scores Before and After Treatment.

**Fig 1:**
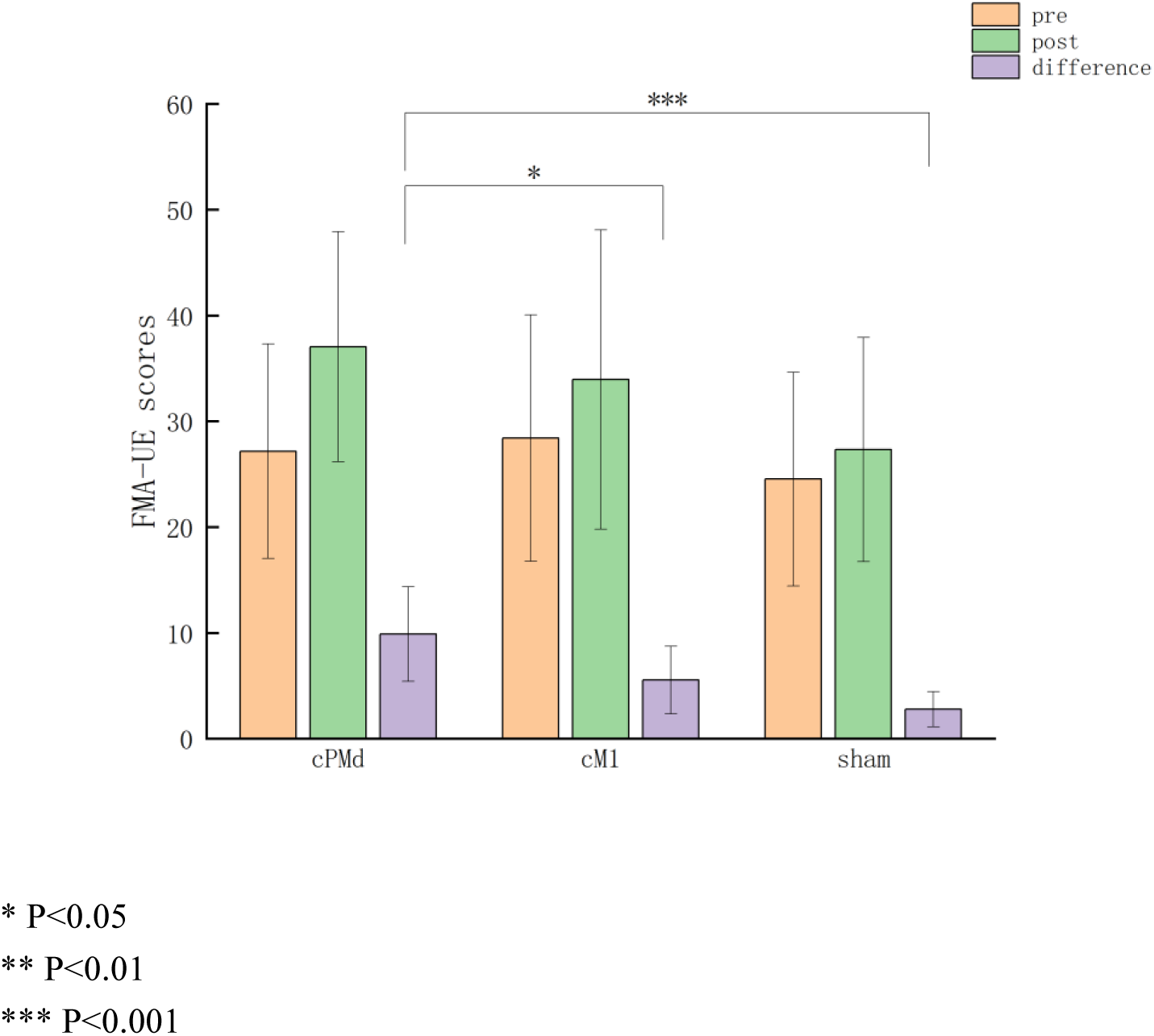
Comparison of FMA-UE scores before and after treatment among three groups

**Fig 2:**
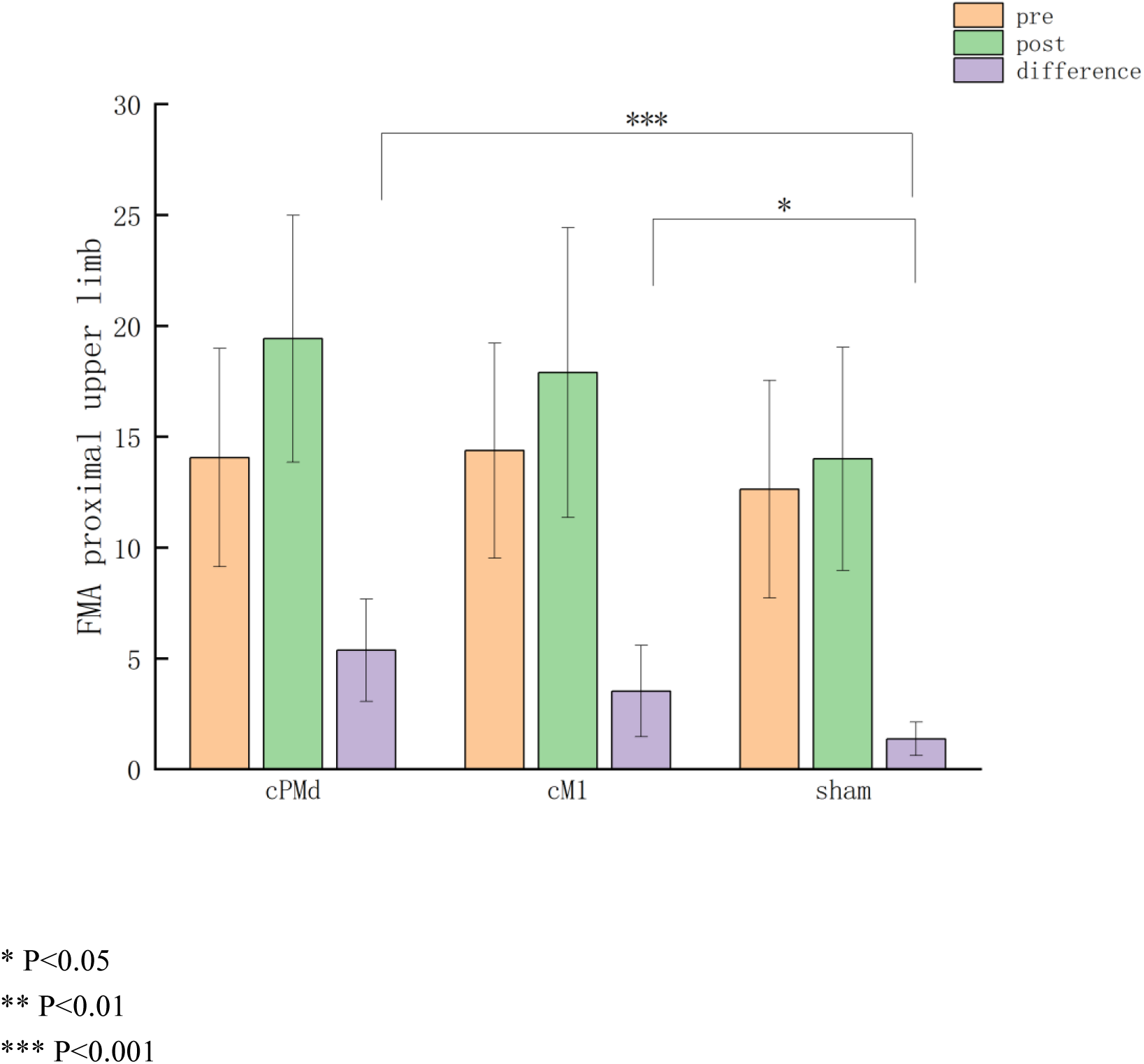
Comparison of FMA proximal upper limb scores before and after treatment among three groups

**Fig 3:**
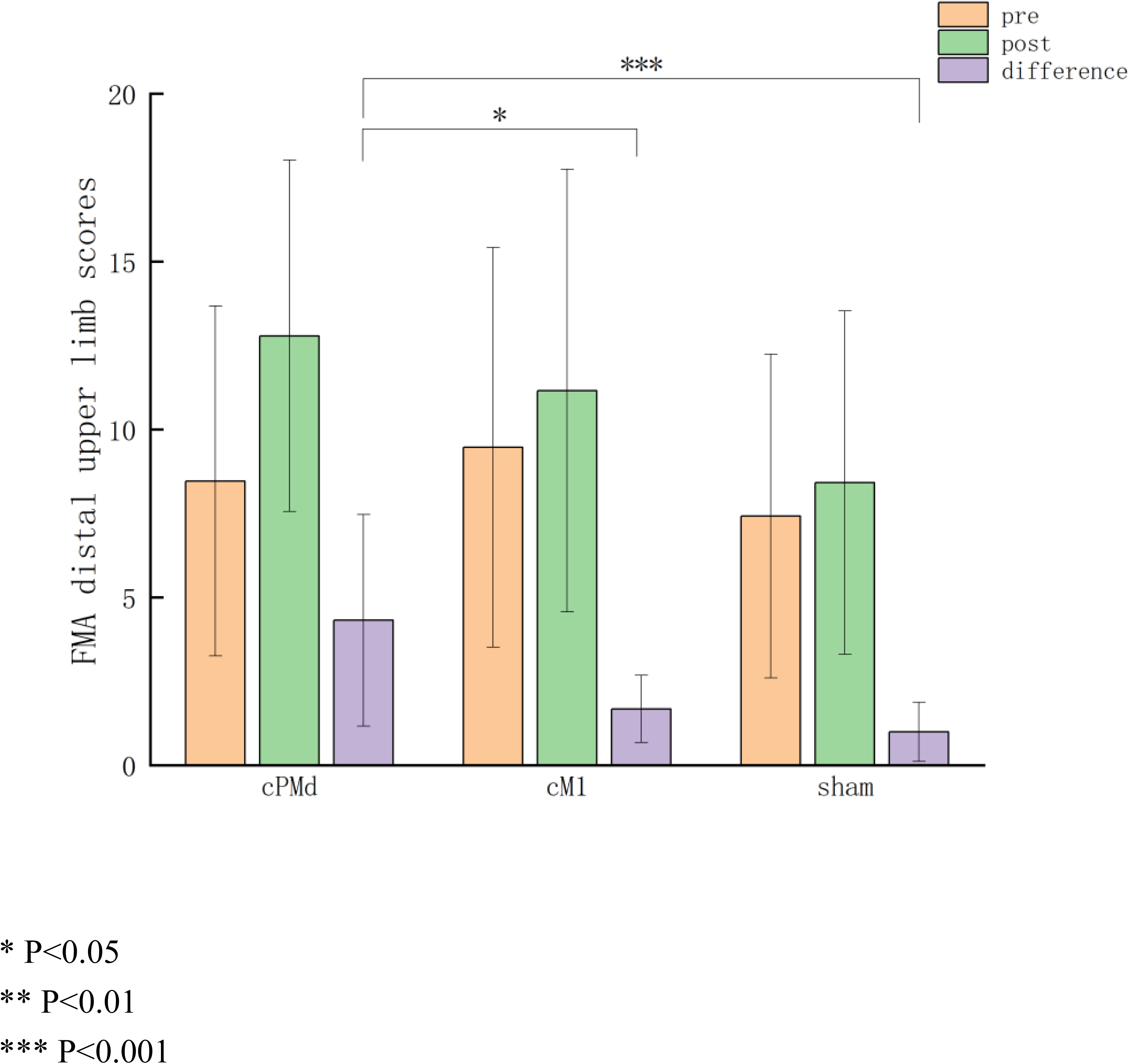
Comparison of FMA distal upper limb scores before and after treatment among three groups

**Fig 4:**
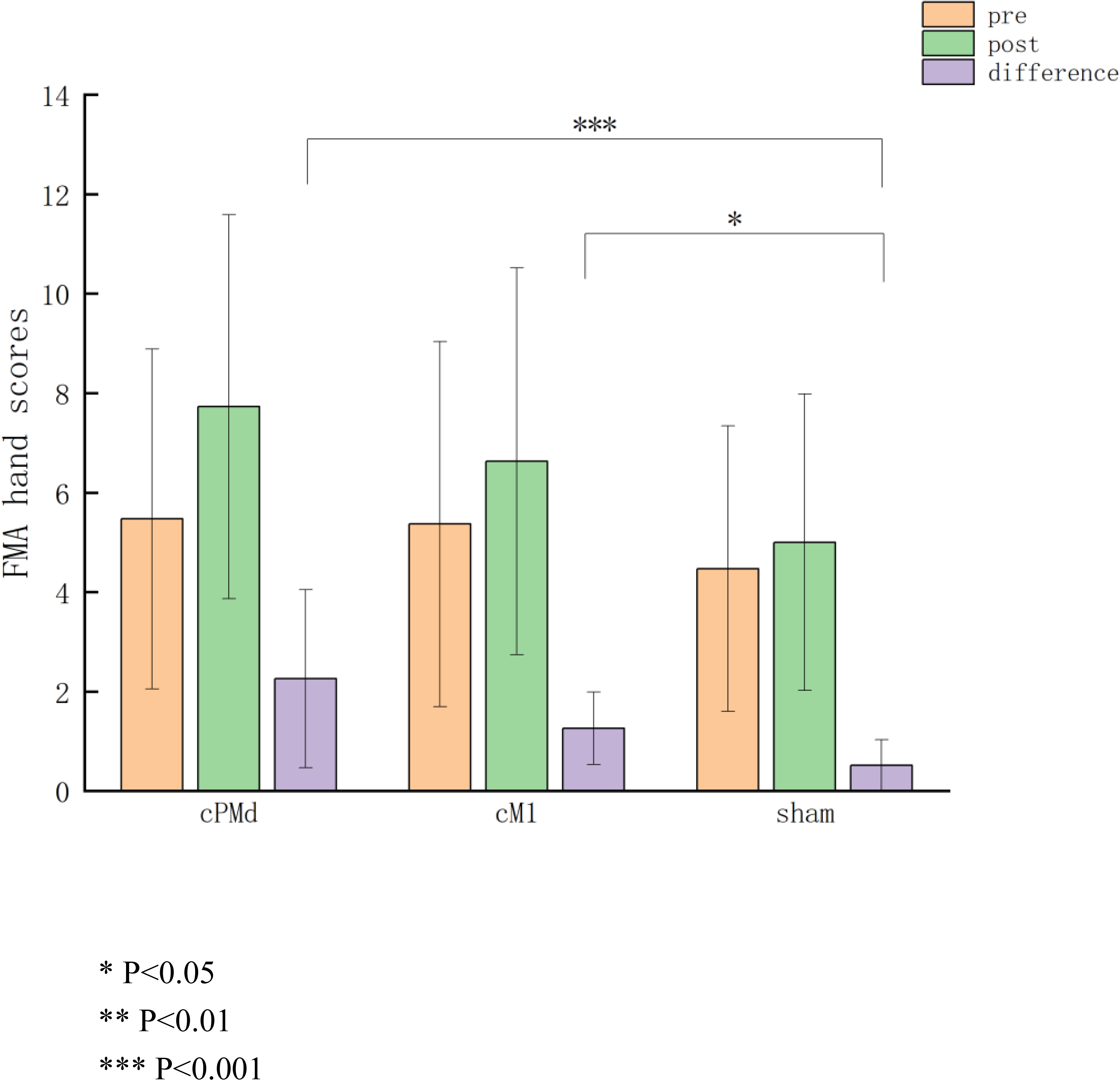
Comparison of FMA hand scores before and after treatment among three groups

### 4. Comparison of FMA Proximal Upper Limb Scores Before and After Treatment

The FMA proximal upper limb scores significantly improved in all three groups post-treatment (P < 0.05). The cPMd group demonstrated significantly greater improvements in FMA proximal upper limb scores compared to the sham group (P < 0.05). Both the cPMd and cM1 groups showed significantly greater improvements compared to the sham group (P < 0.05), although no significant difference was observed between the cPMd and cM1 groups (P > 0.05), as detailed in Table 5.

**Table 5:** Comparison of FMA Proximal Upper Limb Scores Before and After Treatment.

| Group | Before treatment | After treatment<sup>1</sup> | Difference<sup>1</sup> |
| --- | --- | --- | --- |
| **cPMd group(n=19)** | 14.05 ± 4.92 | 19.42 ± 5.57<sup>2ab</sup> | 5.37 ± 2.31<sup>b</sup> |
| **cM1 group(n=19)** | 14.37 ± 4.84 | 17.89 ± 6.53<sup>2a</sup> | 3.52 ± 2.06<sup>b</sup> |
| **sham group (n=19)** | 12.63 ± 4.90 | 14.00 ± 5.03<sup>2a</sup> | 1.37 ± 0.76 |

<sup>1</sup>Due to significant deviations from normality (P<0.05), the Kruskal-Wallis H test was used for between-group comparisons.
<sup>2</sup> Due to significant deviations of difference from normality (P<0.05), the Wilcoxon signed-rank tests were used for pre-post comparisons.
<sup>a</sup>P< 0.05, indicates a statistically significant difference compared to before treatment within the
group;
<sup>b</sup> $P < 0.05$ , indicates a statistically significant difference compared to the control;
<sup>c</sup> $P < 0.05$ , indicates a statistically significant difference compared to the M1 group.

### 5. Comparison of FMA Distal Upper Limb Scores Before and After Treatment

The FMA distal upper limb scores significantly improved in all three groups post-treatment (P < 0.05), although no significant differences were observed among the groups (P > 0.05). The cPMd group exhibited significantly greater improvements compared to both the sham and cM1 groups (P < 0.05), as detailed in Table 6.

**Table 6:** Comparison of FMA Distal Upper Limb Scores Before and After Treatment.

| Group | Before treatment | After treatment<sup>1</sup> | Difference<sup>1</sup> |
| --- | --- | --- | --- |
| **cPMd group(n=19)** | 8.47 ± 5.21 | 12.79 ± 5.23<sup>2a</sup> | 4.32 ± 3.15<sup>bc</sup> |
| **cM1 group(n=19)** | 9.47 ± 5.95 | 11.16 ± 6.59<sup>2a</sup> | 1.68 ± 1.00 |
| **sham group (n=19)** | 7.42 ± 4.82 | 8.42 ± 5.11<sup>2a</sup> | 1.00 ± 0.88 |

<sup>1</sup>Due to significant deviations from normality ( $P < 0.05$ ), the Kruskal-Wallis H test was used for between-group comparisons.
<sup>2</sup> Due to significant deviations of difference from normality ( $P < 0.05$ ), the Wilcoxon signed-rank tests were used for pre-post comparisons.
<sup>a</sup> $P < 0.05$ , indicates a statistically significant difference compared to before treatment within the group;
<sup>b</sup> $P < 0.05$ , indicates a statistically significant difference compared to the control;
<sup>c</sup> $P < 0.05$ , indicates a statistically significant difference compared to the M1 group.

### 6. Comparison of FMA Hand Scores Before and After Treatment

Post-treatment analysis revealed significant improvements in FMA hand scores across all three patient groups (P<0.05), with no statistically significant intergroup differences observed (P>0.05). Comparative analysis demonstrated that both the cPMd and cM1 groups exhibited significantly greater improvements compared to the sham group (P<0.05), although no statistically significant difference was observed between the cPMd and cM1 groups (P>0.05), as detailed in Table 7.

**Table 7:** Comparison of FMA Hand Scores Before and After Treatment.

| Group | Before treatment | After treatment | Difference<sup>1</sup> |
| --- | --- | --- | --- |
| cPMd group(n=19) | $5.47 \pm 3.42$ | $7.73 \pm 3.86^{2a}$ | $2.26 \pm 1.79^b$ |
| cM1 group(n=19) | $5.37 \pm 3.67$ | $6.63 \pm 3.89^{2a}$ | $1.26 \pm 0.73^b$ |
| sham group (n=19) | $4.47 \pm 2.87$ | $5.00 \pm 2.98^{2a}$ | $0.52 \pm 0.51$ |

<sup>1</sup>Due to significant deviations from normality ( $P < 0.05$ ), the Kruskal-Wallis H test was used for between-group comparisons.
<sup>2</sup> Due to significant deviations of difference from normality ( $P < 0.05$ ), the Wilcoxon signed-rank tests were used for pre-post comparisons.
<sup>a</sup> $P < 0.05$ , indicates a statistically significant difference compared to before treatment within the group;
<sup>b</sup> $P < 0.05$ , indicates a statistically significant difference compared to the control;
<sup>c</sup> $P < 0.05$ , indicates a statistically significant difference compared to the M1 group.

### 7. Comparison of FMA-LE Scores Before and After Treatment

Post-treatment analysis demonstrated significant improvements in FMA-LB scores across all three groups (P<0.05). The cPMd group had a difference in scores after treatment compared to the control group (P<0.05). Notably, the cPMd group demonstrated significantly greater score improvements than the sham group (P<0.05). While the cM1 group showed greater improvement than the sham group, this difference did not reach statistical significance (P=0.051), as detailed in Table 8.

**Table 8:** Comparison of FMA-LE Scores Before and After Treatment.

| Group | Before treatment | After treatment<sup>1</sup> | Difference<sup>1</sup> |
| --- | --- | --- | --- |
| **cPMd group(n=19)** | 22.32 ± 3.53 | 28.05 ± 3.49<sup>ab</sup> | 5.74 ± 4.00<sup>b</sup> |
| **cM1 group(n=19)** | 20.89 ± 4.07 | 25.37 ± 3.70<sup>a</sup> | 4.47 ± 2.17 |
| **sham group (n=19)** | 21.00 ± 5.31 | 23.58 ± 5.60<sup>a</sup> | 2.58 ± 1.43 |

<sup>1</sup>Due to heterogeneity of variance (P< 0.05), the Kruskal-Wallis H tests were used for group comparisons.
<sup>a</sup>P< 0.05, indicates a statistically significant difference compared to before treatment within the group;
<sup>b</sup>P< 0.05, indicates a statistically significant difference compared to the sham group;
<sup>c</sup>P< 0.05, indicates a statistically significant difference compared to the cM1 group.

### 8 Comparison of ARAT Scores Before and After Treatment

Post-treatment analysis revealed significant improvements in ARAT scores across all three patient groups (P<0.05), with no statistically significant intergroup differences observed (P>0.05). Further analysis confirmed the absence of significant improvement differences among the three groups (P>0.05), as detailed in Table 9.

**Table 9:** Comparison of ARAT Scores Before and After Treatment of Three Groups.

| Group | Before treatment<sup>1</sup> | After treatment<sup>1</sup> | Difference<sup>1</sup> |
| --- | --- | --- | --- |
| cPMd group(n=19) | 8.94 ± 9.19 | 14.26 ± 12.14<sup>2a</sup> | 5.32 ± 4.58 |
| cM1 group(n=19) | 13.37 ± 11.09 | 16.63 ± 13.12<sup>2a</sup> | 3.26 ± 2.64 |
| sham group (n=19) | 9.16 ± 7.11 | 12.63 ± 8.78<sup>2a</sup> | 3.47 ± 2.80 |

<sup>1</sup>Due to significant deviations from normality (P< 0.05), the Kruskal-Wallis H test was used for between-group comparisons.
<sup>2</sup> Due to significant deviations of difference from normality (P<0.05), the Wilcoxon signed-rank tests were used for pre-post comparisons.
<sup>a</sup>P< 0.05, indicates a statistically significant difference compared to before treatment within the group;
<sup>b</sup>P< 0.05, indicates a statistically significant difference compared to the sham group;
<sup>c</sup>P< 0.05, indicates a statistically significant difference compared to the cM1 group.

### 9. Comparison of WMFT Scores Before and After Treatment

Post-treatment analysis demonstrated significant improvements in WMFT scores across all three patient groups (P<0.05), with no statistically significant intergroup differences observed (P>0.05). Further comparative analysis confirmed the absence of significant improvement differences among the three groups (P>0.05), as detailed in Table 10.

**Table 10:** Comparison of WMFT Scores Before and After Treatment among Three Groups.

| Group | Before treatment<sup>1</sup> | After treatment<sup>1</sup> | Difference<sup>1</sup> |
| --- | --- | --- | --- |
| cPMd group(n=19) | 14.63 ± 11.96 | 23.74 ± 18.41<sup>2a</sup> | 9.10 ± 10.14 |
| cM1 group(n=19) | 10.05 ± 8.98 | 13.05 ± 10.08<sup>2a</sup> | 3.00 ± 1.80 |
| sham group (n=19) | 9.95 ± 9.05 | 13.89 ± 10.91<sup>2a</sup> | 3.94 ± 3.37 |

<sup>1</sup>Due to significant deviations from normality (p < 0.05), the Kruskal-Wallis H test was used for between-group comparisons.
<sup>2</sup> Due to significant deviations of difference from normality (p < 0.05), the Wilcoxon signed-rank tests were used for pre-post comparisons.
<sup>a</sup>P< 0.05, indicates a statistically significant difference compared to before treatment within the group;
<sup>b</sup>P< 0.05, indicates a statistically significant difference compared to the sham group;
<sup>c</sup>P< 0.05, indicates a statistically significant difference compared to the cM1 group.

### 10. Comparison of BI Scores Before and After Treatment

BI scores of the three groups were significantly increased before and after treatment (P<0.05), and cPMd group was significantly different from control group and cM1 after treatment (P<0.05). The improvement of the cPMd group and the cM1 group was significantly greater than the sham group (P<0.05), but the difference between two groups was not statistically significant (P>0.05), as shown in Table 11.

**Table 11:** Comparison of BI Scores Before and After Treatment among the Three Groups.

| Group | Before treatment | After treatment | Difference |
| --- | --- | --- | --- |
| cPMd group(n=19) | 64.50± 15.46 | 80.25±11.97<sup>ab</sup> | 15.75±10.91<sup>b</sup> |
| cM1 group(n=19) | 60.50± 14.94 | 73.00±14.17<sup>a</sup> | 12.50±5.25<sup>b</sup> |
| sham group (n=19) | 56.50± 10.64 | 64.50±9.58<sup>a</sup> | 8.00±2.99 |

<sup>a</sup> $P<0.05$ , indicates a statistically significant difference compared to before treatment within the group;
<sup>b</sup> $P<0.05$ , indicates a statistically significant difference compared to the sham group;
<sup>c</sup> $P<0.05$ , indicates a statistically significant difference compared to the cM1 group.

### 11. Comparison of MEP Amplitude Before and After Treatment

Post-treatment analysis demonstrated significant increases in ipsilesional MEP amplitude across all three patient groups (P<0.05), with no statistically significant intergroup differences observed (P>0.05). Further comparative analysis confirmed the absence of significant improvement differences among the three groups (P>0.05), as detailed in Table 12.

**Table 12:** Comparison of Ipsilesional MEP Amplitude on the Affected Side Before and After Treatment among the Three Groups.

| Group | Before treatment | After treatment | Difference<sup>1</sup> |
| --- | --- | --- | --- |
| cPMd group(n=10/19) | 217.60± 58.14 | 231.50± 54.86<sup>2a</sup> | 13.90± 27.20 |
| cM1 group(n=9/19) | 222.56± 76.28 | 235.78± 75.86<sup>2a</sup> | 13.22± 10.07 |
| sham group (n=10/19) | 219.60± 72.78 | 235.10± 72.92<sup>a</sup> | 15.50± 13.88 |

<sup>1</sup>Due to significant deviations from normality ( $P<0.05$ ), the Kruskal-Wallis H test was used for between-group comparisons.
<sup>2</sup> Due to significant deviations of difference from normality ( $P<0.05$ ), the Wilcoxon signed-rank tests were used for pre-post comparisons.
<sup>a</sup> $P < 0.05$ , indicates a statistically significant difference compared to before treatment within the group;
<sup>b</sup> $P < 0.05$ , indicates a statistically significant difference compared to the sham group;
<sup>c</sup> $P < 0.05$ , indicates a statistically significant difference compared to the cM1 group.

### 12. Comparison of MEP Latency Before and After Treatment

Post-treatment analysis demonstrated significant shortening of ipsilesional MEP latency in both the cPMd and cM1 groups (P<0.05). However, comparative analysis revealed no statistically significant improvement differences among the three groups (P>0.05), as detailed in Table 13.

**Table 13:** Comparison of Ipsilesional MEP Latency Before and After Treatment among the Three Groups.

| Group | Before treatment | After treatment<sup>1</sup> | Difference<sup>1</sup> |
| --- | --- | --- | --- |
| cPMd group(n=10/19) | 22.64 ± 0.60 | 22.20 ± 0.61<sup>a</sup> | -0.43 ± 0.31 |
| cM1 group(n=9/19) | 23.03 ± 0.97 | 22.72 ± 0.80<sup>a</sup> | -0.32 ± 0.36 |
| sham group (n=10/19) | 22.96 ± 0.70 | 22.86 ± 0.61<sup>2</sup> | -0.1 ± 0.31 |

<sup>1</sup>Due to significant deviations from normality ( $P < 0.05$ ), the Kruskal-Wallis H test was used for between-group comparisons.
<sup>2</sup>Due to significant deviations of difference from normality ( $P < 0.05$ ), the Wilcoxon signed-rank tests were used for pre-post comparisons.
<sup>a</sup> $P < 0.05$ , indicates a statistically significant difference compared to before treatment within the group;
<sup>b</sup> $P < 0.05$ , indicates a statistically significant difference compared to the sham group;
<sup>c</sup> $P < 0.05$ , indicates a statistically significant difference compared to the cM1 group.

### 13. Comparison of Ipsilesional Central Conduction Time Before and After Treatment

Post-treatment analysis revealed significant shortening of ipsilesional central conduction time across all three groups (P<0.05), with no statistically significant intergroup differences observed (P>0.05). Notably, the cPMd group demonstrated significantly greater improvement compared to the sham group (P<0.05), as detailed in Table 14.

**Table 14:**
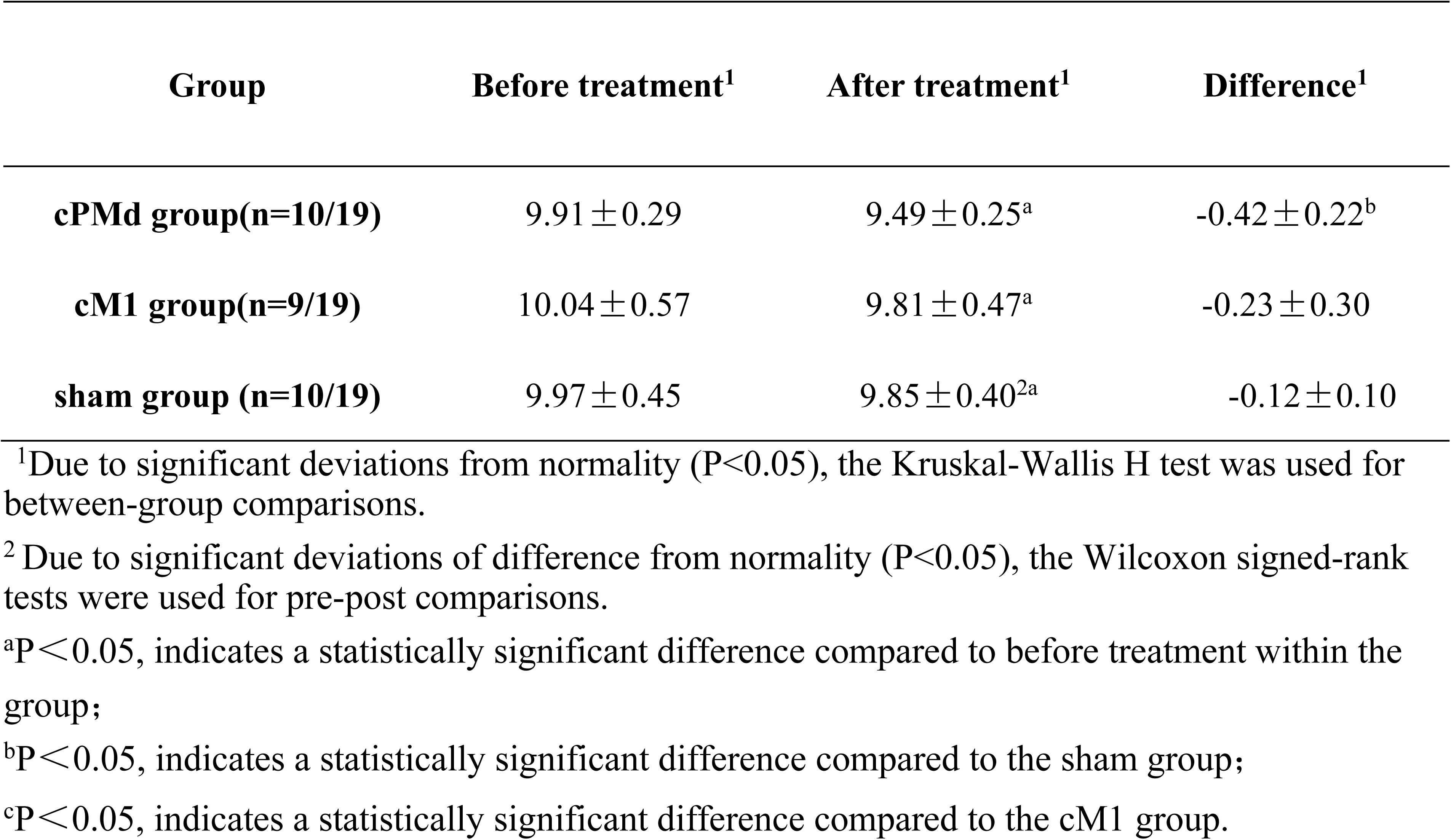
Comparison of Ipsilesional Central Conduction Time Before and After Treatment among the Three Groups.

### 14. Changes of MEP Amplitude on the Healthy Side Before and After Treatment

Post-treatment analysis demonstrated a significant increase in MEP amplitude on the healthy side in the cPMd group (P<0.05). Comparative analysis revealed that both the cPMd and sham groups exhibited significantly greater improvements in MEP amplitude compared to the cM1 group (P<0.05), as detailed in Table 15.

**Table 15:**
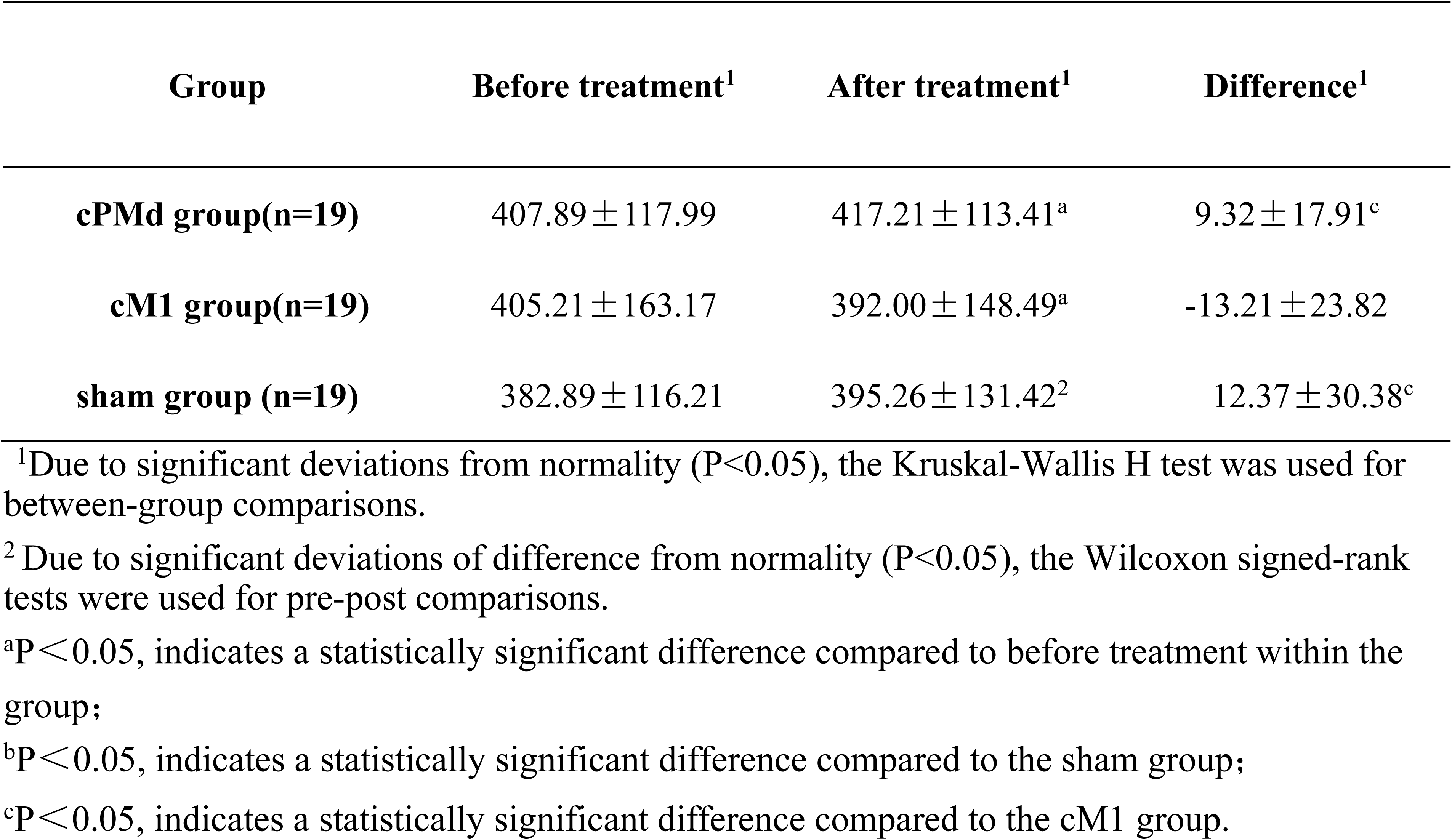
Comparison of MEP Amplitude on the Healthy Side Before and After Treatment among the Three Groups.

## Discussion

Transcranial magnetic stimulation (TMS), a non-invasive neuromodulation technique, has demonstrated significant clinical efficacy in enhancing upper limb motor function recovery in stroke patients. TMS effectively enhances neural plasticity, restores cortical function, and improves upper limb motor performance. Currently, the development of transcranial magnetic stimulation (TMS) treatment protocols in clinical practice is predominantly guided by the interhemispheric inhibition theory, which involves either the excitation of ipsilesional motor cortex activity or the inhibition of contralesional motor cortex activity. However, the therapeutic efficacy of these protocols is contingent upon the severity of the stroke and the integrity of the corticospinal tract[17]. Consequently, Di Pino et al. introduced the bimodal balance–recovery model in 2014, which provides a more nuanced framework for understanding post-stroke recovery[18]. Emerging evidence suggests that the contralesional premotor dorsal area (cPMd) may serve as a promising stimulation target[10, 11]. Anatomically, the fibers originating from the PMd region project directly to the brainstem and exhibit robust connectivity with the ipsilateral upper limb, potentially providing the structural foundation for cPMd-mediated compensation of motor function on the affected side[19]. Behavioral studies further corroborate the compensatory role of PMd in ipsilateral upper limb movement. For instance, in healthy individuals performing specific hand movement tasks, inhibition of cPMd activity did not impair task performance, while ipsilesional PMd excitability increased. Notably, when dual-pulse transcranial magnetic stimulation (DP-TMS) transiently suppressed ipsilesional PMd activity, task execution errors were observed[20]. These findings suggest that when cPMd activity is inhibited, the ipsilateral hand demonstrates adaptive compensatory mechanisms. Additional evidence indicates that cPMd may also facilitate compensatory engagement in upper limb and finger movement tasks among stroke patients[21].

Our study provides preliminary evidence supporting the efficacy of high-frequency repetitive transcranial magnetic stimulation (rTMS) targeting the cPMd in ameliorating post-stroke motor impairments. Comparative analysis revealed that the cPMd group demonstrated significant improvements in both the Fugl-Meyer Assessment for Upper Extremity (FMA-UE) and distal upper limb scores relative to the cM1 group. Furthermore, the cPMd group exhibited marked enhancements in FMA for Lower Extremity (FMA-LE), proximal upper limb, and hand scores compared to the sham stimulation group. These observed improvements may be attributed to the PMd’s role as a motor planning region, which integrates sensory inputs, coordinates multi-joint sequential movements, and regulates the timing of complex motor actions[16]. The PMd’s extensive connectivity with the posterior parietal lobe and cerebellum may directly contribute to the encoding of distal fine motor skills, including independent finger flexion and extension, as well as grip force modulation. However, the study revealed limited improvements in electrophysiological assessments, with only ipsilesional central conduction time showing significant changes compared to the sham group. This limitation may be partially explained by the restricted sample size, as motor evoked potentials (MEPs) were elicitable in only approximately 50% of participants across all groups. These findings collectively suggest that high-frequency rTMS targeting the cPMd may represent a novel and potentially more effective approach for optimizing motor network reorganization and improving functional deficits in stroke patients. This study offers new perspectives for TMS-based assessment and intervention strategies grounded in complex brain network analysis, while proposing innovative therapeutic approaches for stroke rehabilitation. Nevertheless, the current investigation is limited by its relatively small sample size and insufficient exploration of underlying mechanisms. Future research should prioritize multicenter, large-scale systematic studies to further investigate the therapeutic potential of high-frequency rTMS targeting the cPMd in stroke rehabilitation. Additionally, comprehensive investigations into the pathophysiological mechanisms underlying stroke recovery and the neural plasticity associated with motor dysfunction rehabilitation are warranted. Consequently, the therapeutic potential of high-frequency rTMS targeting the cPMd requires further exploration to optimize its application in promoting motor function recovery in stroke patients.

## Data Availability

All data produced in the present study are available upon reasonable request to the authors

